# Meteorological Influence on Atrial Fibrillation and Flutter, A Nationwide Observational Study

**DOI:** 10.1101/2023.07.11.23292530

**Authors:** Andrew Geunwon Kim, Chanjoo Park, Nithi Tokavanich, Rand Sabanci, Rebeccah Freel, Victoria Hayes, Ranjan Thakur

**Affiliations:** Department of Internal Medicine, Michigan State University, East Lansing, MI, United States; Department of Medicine, Catholic Kwandong University International St. Mary’s Hospital, Incheon, South Korea; Department of Cardiac Electrophysiology, University of Michigan, Ann Arbor, MI, United States; Department of Cardiac Electrophysiology, Thoracic & Cardiovascular Institute, Sparrow Hospital, Lansing, MI, United States

**Keywords:** Categories, Special sections: Atrial fibrillation, atrial flutter, meteorology, climate, weather, air pollutants, particulate matter, nitrogen dioxide, sulfur dioxide, ozone, carbon monoxide, heat, temperature, sunlight, humidity, precipitation, wind speed, incidence, prevalence, arrhythmia

## Abstract

**Background:** The impact of meteorological factors, including atmospheric temperature, humidity, wind speed, and others, on the incidence of atrial fibrillation and flutter (AF) has been the subject of several studies, but the findings have been inconsistent. Given the complex and multifaceted nature of this relationship, a larger-scale study was necessary to provide sufficient statistical power and elucidate potential associations between them.

**Methods:** The South Korean government provides open access to national health insurance and weather data for its more than 50 million citizens from January 2010 to July 2022. The national health insurance data includes the monthly number of patients diagnosed with a specific condition, reflecting the incidence and prevalence of the condition. Pearson correlation analyses were performed using the statistical analysis software SAS for Academics to examine the association between each month’s national average climate data and the number of patients diagnosed with AF.

**Results:** The number of patients diagnosed with AF in the total population showed a statistically significant correlation only with average wind speed (r=-0.42, 95% CI -0.55 to -0.28, p<0.001) and sunshine duration (r=0.27, 95% CI 0.12 to 0.41, p<0.001). Among females aged 20 to 24 years, there was a statistically significant association with other variables, including average temperature, precipitation, humidity, and atmospheric pressure (p<0.05). Diurnal temperature variation showed inconsistent associations across different age and sex groups.

**Conclusions:** The number of patients diagnosed with AF is negatively correlated with average wind speed and positively correlated with sunshine duration in the general population, particularly among the elderly. There was no significant association between the number of patients diagnosed with AF and average temperature, precipitation, or humidity, except for females aged 20 to 24 years, who exhibited a significant association with these variables. However, it is important to note that these correlations do not establish causality.

## Introduction

The incidence of atrial fibrillation and flutter (AF) has been extensively studied in relation to meteorological factors, including atmospheric temperature, humidity, wind speed, and others. However, these studies have yielded inconsistent findings, necessitating a larger-scale investigation to enhance statistical power and clarify the potential associations between meteorological factors and AF.

Temperature variability has been linked to the risk of cardiac arrhythmias, including AF [1]. Nonetheless, the significance of atmospheric temperatures in relation to AF remains inconclusive [19, 20]. Several studies have suggested that low temperatures, rather than high temperatures, increase the risk of AF [2-9]. Colder temperatures are believed to impact microvascular function and resistance, placing strain on the cardiovascular system [10]. Another theory proposes that heat-induced heat shock proteins may offer protection against myocardial remodeling and the development of AF in higher temperatures [11, 12].

However, it is important to note that alcohol consumption plays a significant confounding role in cold weather, as it is a well-known major risk factor for AF and alcohol sales tend to increase during colder seasons [13]. On the other hand, certain studies indicate that high temperatures may also elevate the risk of atrial fibrillation, particularly in the presence of heat stroke risk factors such as high temperature, high humidity, precipitation, and prolonged sunlight exposure [14, 15, 18]. Overall, it appears that both extreme cold and hot temperatures increase the risk of AF [16-18].

Other meteorological factors, including humidity, precipitation, wind speed, volatile gases, and fine particles in the air, have also been examined, but the results have been inconsistent. The effect of humidity and precipitation on the risk of AF has been reported as increasing [14, 15, 18], decreasing [8], or having no significant effect [4, 19, 21]. Fine particles, such as particulate matter with a diameter of 2.5 micrometers or smaller (PM_2.5_), as well as PM_10_, have demonstrated an increased risk in certain studies [3, 9, 24, 26, 27], while others did not find a statistically significant association with PM_2.5_ [28] or PM_10_ [25]. Associations have been observed between elevated levels of other air pollutants, such as ozone (O_3_) [23], nitrogen dioxide (NO_2_) [7, 24, 25], and sulfur dioxide (SO_2_) [24], and the risk of AF. However, some studies did not find a statistically significant association with O_3_ [24, 27], NO_2_ [27], or SO_2_ [27]. The relationship between carbon monoxide (CO) and AF has been inconsistent, with some studies reporting an association [9] and others finding no statistical significance [24, 27].

Atmospheric pressure and wind speed significantly influence the concentration and dispersion of air pollutants [22]. Increased atmospheric pressure has been correlated with a higher incidence of AF [18, 21]. Low wind speeds can result in the stagnation of air pollutants, leading to elevated levels of particulate matter and nitrogen dioxide, which are known potential contributors to AF [22]. However, different studies have produced conflicting results. One study suggests that high wind speeds, rather than low wind speeds, are associated with an increased incidence of AF [9], while another study reports no significant effect of wind speed on AF [19].

Additionally, it is worth noting that the impact of meteorological factors on humans may be diminishing [19] due to the increasing amount of time spent indoors and the advancements in air conditioning systems, which have effectively reduced the influence of weather on individuals [30].

## Methods

### Healthcare data

The Healthcare Big Data Hub, operated by the South Korean government, provides open access to national health insurance data (opendata.hira.or.kr) for approximately 50 million citizens. This comprehensive dataset includes information on the monthly number of patients diagnosed with specific conditions, categorized according to the Korean Standard Classification of Diseases (KCD). The KCD system is a slightly modified version of the International Classification of Diseases 10 (ICD-10). The data is further segmented by age, in 5-year intervals, and sex groups, allowing for detailed analysis. The available data covers the period from January 2010 to July 2022.

### Meteorological data

The Korea Meteorological Association (kma.go.kr) offers open access to national climate data, including metrics such as national average temperature, precipitation, humidity, wind speed, and sunshine hours. Additional weather indices, such as the heat index and wind chill index, were calculated using formulas from the United States National Weather Service (weather.gov). Environmental and air pollution data were obtained from the Korea Statistical Information Service (kosis.kr).

### Statistical analysis

Pearson correlation analyses were conducted using the statistical analysis software SAS for Academics to examine the potential associations between the national average climate data for each month and the number of patients diagnosed with atrial fibrillation or flutter.

## Results

### Healthcare data [Figure 1, Table 1]

In 2010, the National Health Insurance registered a total population of approximately 48.9 million people, consisting of 24.6 million males and 24.3 million females. By 2021, the total population had increased to about 51.4 million, with 25.7 million males and 25.7 million females. Over time, there has been a significant advancement in the age distribution of the population. The median age was 38.1 in 2010 and 44.5 in 2021, while the mean age was 38.1 in 2010 and 43.5 in 2021. Baseline data for 2022 is currently unavailable. Although the total population has not experienced significant growth, there has been a notable increase in the number of patients diagnosed with AF over the years.

**Table 1.** Baseline characteristics of the total population

|  | 2010 | 2021 |
| --- | --- | --- |
| Total Population | 48,906,765 | 51,412,137 |
| Male | 24,648,532 | 25,742,882 |
| Female | 24,258,263 | 25,669,255 |
| Male-to-Female Ratio | 1.016 | 1.003 |
| Mean Age | 38.1 | 43.5 |
| Median Age | 38.1 | 44.5 |

**Figure 1.**
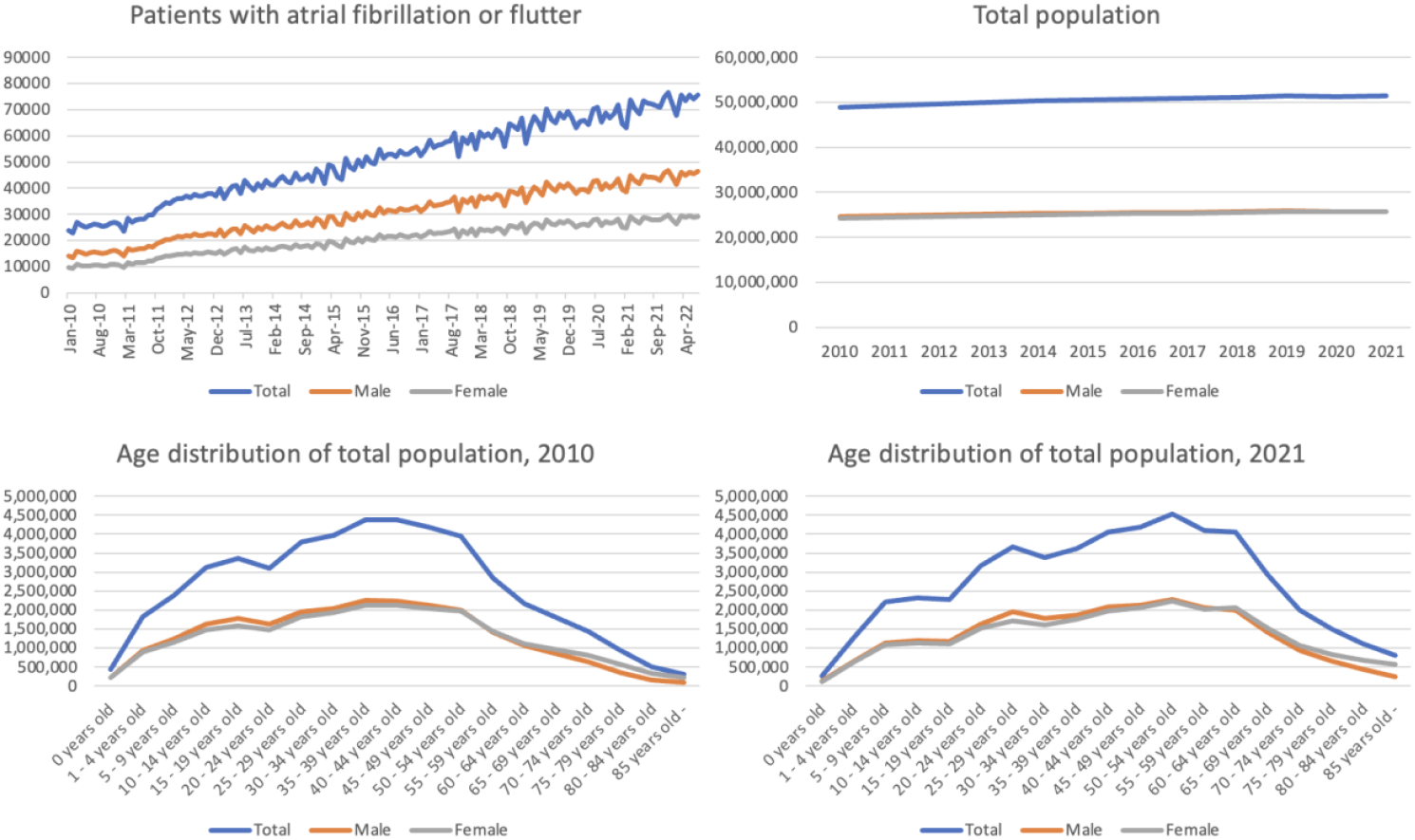
Baseline characteristics of total population and the number of patients with AF

### Meteorological data [Figure 2, Table 2]

South Korea features a continental climate characterized by very cold, dry winters and very hot, humid summers. Spring and autumn are relatively short, and temperatures are mild and generally quite pleasant. There are only minimal regional variations in weather conditions throughout the country. Most air pollutants have been steadily reduced over the years following green energy policy. Of note, weather indices are intended for certain weather conditions. For example, the wind chill index is applied in winter, while the heat index is applied in summer.

**Table 2.**
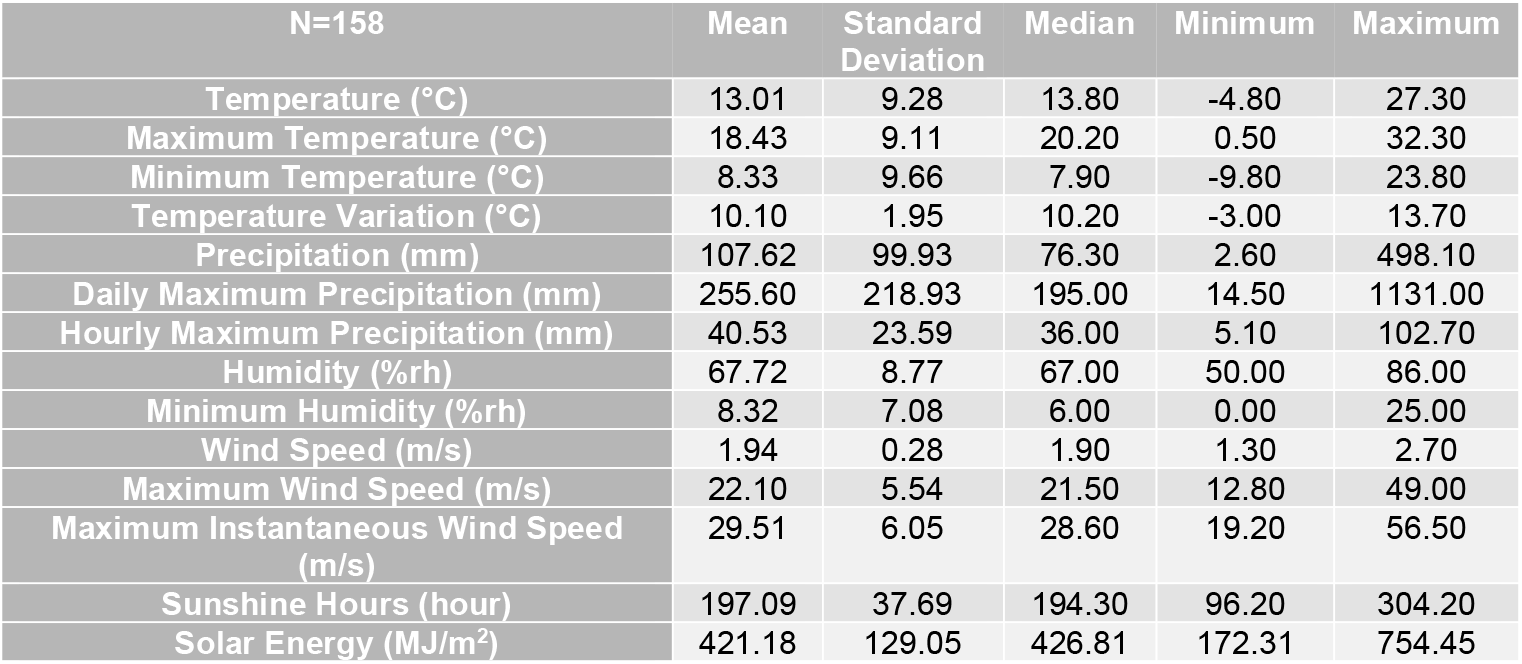
Simple statistics of the weather variables

| N=158 | Mean | Standard Deviation | Median | Minimum | Maximum |
| --- | --- | --- | --- | --- | --- |
| Temperature (°C) | 13.01 | 9.28 | 13.80 | -4.80 | 27.30 |
| Maximum Temperature (°C) | 18.43 | 9.11 | 20.20 | 0.50 | 32.30 |
| Minimum Temperature (°C) | 8.33 | 9.66 | 7.90 | -9.80 | 23.80 |
| Temperature Variation (°C) | 10.10 | 1.95 | 10.20 | -3.00 | 13.70 |
| Precipitation (mm) | 107.62 | 99.93 | 76.30 | 2.60 | 498.10 |
| Daily Maximum Precipitation (mm) | 255.60 | 218.93 | 195.00 | 14.50 | 1131.00 |
| Hourly Maximum Precipitation (mm) | 40.53 | 23.59 | 36.00 | 5.10 | 102.70 |
| Humidity (%rh) | 67.72 | 8.77 | 67.00 | 50.00 | 86.00 |
| Minimum Humidity (%rh) | 8.32 | 7.08 | 6.00 | 0.00 | 25.00 |
| Wind Speed (m/s) | 1.94 | 0.28 | 1.90 | 1.30 | 2.70 |
| Maximum Wind Speed (m/s) | 22.10 | 5.54 | 21.50 | 12.80 | 49.00 |
| Maximum Instantaneous Wind Speed (m/s) | 29.51 | 6.05 | 28.60 | 19.20 | 56.50 |
| Sunshine Hours (hour) | 197.09 | 37.69 | 194.30 | 96.20 | 304.20 |
| Solar Energy (MJ/m<sup>2</sup>) | 421.18 | 129.05 | 426.81 | 172.31 | 754.45 |

**Figure 2.**
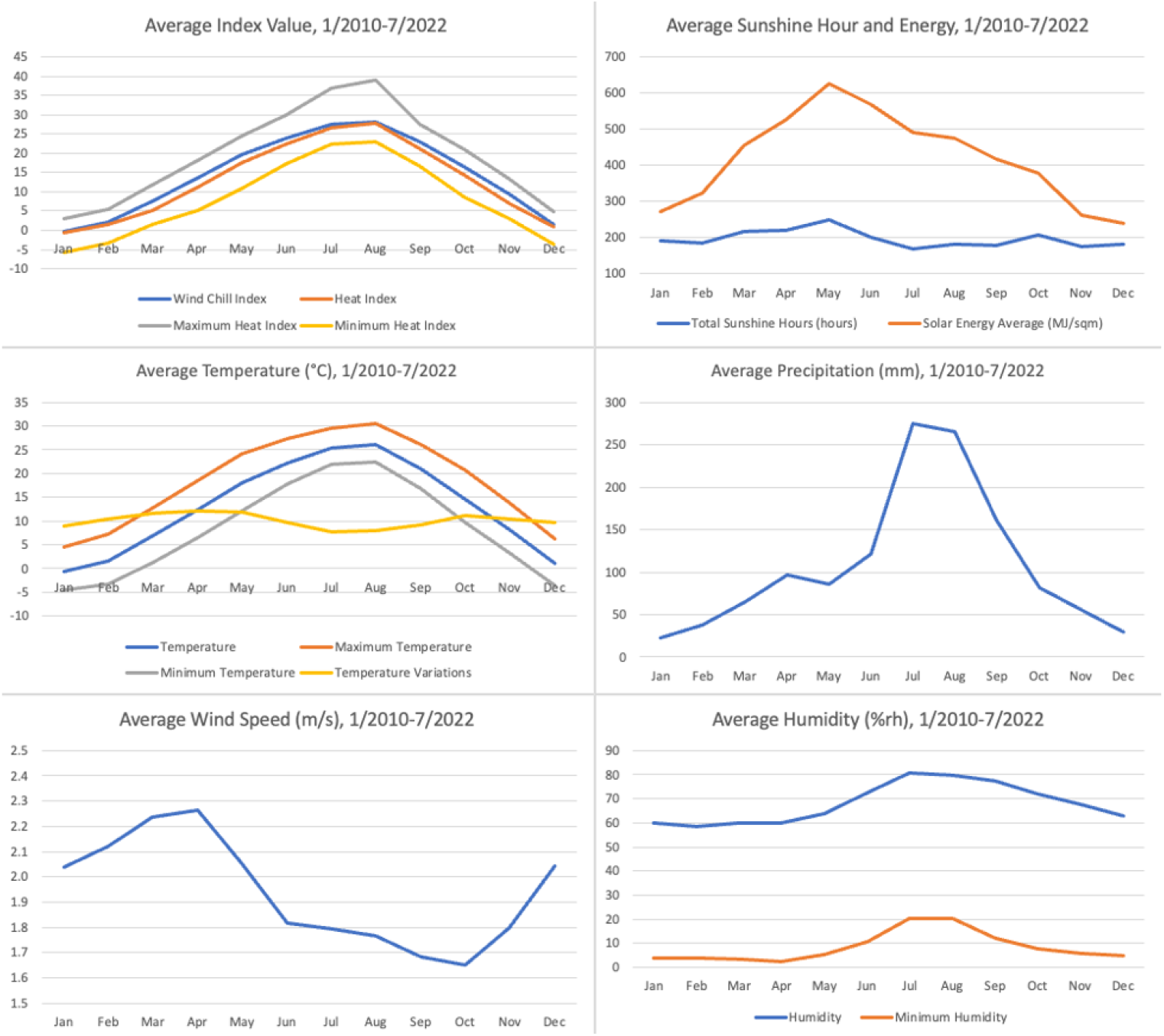
Weather variables

### Statistical analysis

In the overall population, a statistically significant correlation was observed between the number of patients diagnosed with AF and average wind speed (r=-0.42, 95% CI -0.55 to -0.28, p<0.001), as well as sunshine duration (r=0.27, 95% CI 0.12 to 0.41, p<0.001). Subgroup analysis revealed that these correlations were more pronounced in older males and females, particularly in males aged over 35 and females aged over 55. Regarding diurnal temperature variation or intraday temperature variation, its significance varied across different age and sex groups, resulting in inconsistent but statistically significant correlations. In our analysis, notable correlations with the traditional risk factors were observed only among females, predominantly aged 20-24 years. Females in this age group showed a statistically significant association with other variables, including average temperature (r=0.22, 95% CI 0.06 to 0.37, p=0.007), precipitation (r=0.22, 95% CI 0.07 to 0.37, p=0.006), humidity (r=0.22, 95% CI 0.06 to 0.37, p=0.006), and atmospheric pressure (r=-0.22, 95% CI -0.39 to -0.04, p=0.014). However, similar associations were not observed in other age and sex groups.

## Discussion

### Negative correlation between wind speed and AF in the general population

The number of patients diagnosed with AF displayed a negative correlation with average wind speed (r = -0.42, 95% CI -0.55 to - 0.28, p < 0.001). Wind speed has a significant impact on the concentration and dispersion of air pollutants [22]. Lower wind speeds can lead to the stagnation of air pollutants, resulting in higher levels of particulate matter and nitrogen dioxide, both of which are known potential contributors to AF [22].

However, it is worth noting that previous studies have suggested a positive association between higher wind speeds and the occurrence of AF, which is opposite to this study’s findings. Researchers have proposed hypotheses suggesting that higher wind speeds may contribute to cardiovascular stress and AF through mechanisms such as atmospheric pressure oscillations [29], as well as wind-induced increases in the intensity of the atmospheric electric field [9]. Nevertheless, it is important to recognize that these connections remain speculative. Further investigation through experimental studies may be necessary to gain a better understanding of these relationships.

### Positive correlation between sunlight duration and AF in the general population

The number of patients diagnosed with AF showed a positive correlation with monthly sunshine duration (r=0.27, 95% CI 0.12 to 0.41, p<0.001). Prolonged exposure to sunlight may also increase the risk of heat exhaustion or heat stroke, potentially triggering AF [31, 32]. However, there are studies suggesting that sunlight may have a protective effect against paroxysms of atrial fibrillation [33, 34]. Previous research has indicated a higher incidence of AF during colder months when sunshine duration is typically shorter, rather than longer. It is important to consider the close relationship between weather variables in specific locations, which can limit the meaningful interpretation of these findings. For instance, sunshine duration is closely associated with temperature and precipitation. It tends to be longer on high-temperature days and shorter on low-precipitation days, but these relationships can vary depending on the location. For example, in South Korea, sunshine duration is shorter on hot days during the summer due to a higher number of rainy days, whereas in Michigan, USA, it is longer on hot summer days. These region-specific relationships impose limitations on the meaningful interpretation of the correlation between sunlight duration and AF.

### Weather susceptibility of the female population aged 20-24 years

Females in the 20-24-year age group exhibited a statistically significant association with traditional risk factors, including average temperature (r=0.22, 95% CI 0.06 to 0.37, p=0.007), precipitation (r=0.22, 95% CI 0.07 to 0.37, p=0.006), humidity (r=0.22, 95% CI 0.06 to 0.37, p=0.006), and atmospheric pressure (r=-0.22, 95% CI -0.39 to - 0.04, p=0.014). Positive correlations with average temperature, precipitation, and humidity can potentially be attributed to conditions commonly experienced by young females. Autonomic dysfunction, postural orthostatic hypotension (POTS) [37], multiple sclerosis [39-41], and various autoimmune diseases are known to be relatively more prevalent in young females. It is noteworthy that POTS [37], MS [39-41], and certain autoimmune diseases [42] tend to worsen in hot and humid conditions. Therefore, it can be hypothesized that stress induced by the exacerbation of these underlying conditions may trigger episodes of atrial fibrillation in young females. While previous studies have shown positive correlations between atmospheric pressure and atrial fibrillation [18, 21], our findings indicate a negative correlation in young females. This might suggest that the change or gradient of atmospheric pressure may have a greater effect on atrial fibrillation than the actual pressure itself. Nevertheless, meaningful interpretation is significantly constrained by the absence of a causal relationship due to the observational study design and the presence of strong connections and multicollinearities among the variables.

## Limitations

### Independence of events and meaningless correlation

The study is retrospective and observational in nature, which limits its ability to establish a cause-and-effect relationship between variables. In the context of an observational study, it is important to recognize that a statistically significant correlation between two variables does not necessarily indicate a meaningful relationship. Each event can be independent and not directly related to each other. For instance, the increase in AF cases may be influenced by the aging population, while the decrease in air pollutants may be attributed to government initiatives focused on implementing renewable energy policies to reduce emissions. Although statistical analysis may reveal a negative correlation between AF and air pollutants, it does not establish a causal relationship between the two. Each of these factors operates independently, with its own distinct underlying reasons. These confounding factors introduce bias and present challenges in establishing a direct cause-and-effect relationship. Thus, caution should be exercised when interpreting the results, as the correlation may not reflect a meaningful association. To determine the reasons behind the association between a specific factor and a particular outcome, experimental studies would be necessary. However, the multicollinearity of meteorological variables acts as a limitation to designing such a study as well.

### Multicollinearity between meteorological variables

Multicollinearity, also known as collinearity, is a phenomenon in which one predictor variable in a multiple regression model can be accurately predicted from the others with a substantial degree of accuracy. For instance, faster winds may disperse airborne dust, potentially resulting in a decrease in particulate matter concentration [22]. The interconnected nature of meteorological variables poses challenges in isolating their individual effects and conducting precise multiple regression analyses [29]. Furthermore, it is crucial to emphasize that the linear relationship between the meteorological variables can significantly vary depending on the specific location, further complicating the interpretation and analysis of their effects.

### Scientific plausibility

This study revealed statistically significant associations between wind speed, sunlight, and the occurrence of AF. Many hypotheses have been proposed as discussed above regarding the impact of various weather variables on cardiovascular stress, including AF, but some of these hypotheses are speculative and lack experimental evidence.

### Nature and accuracy of data

The data consists of the monthly number of patients recorded for a specific diagnosis under national health insurance. Although it is expected to provide insights into the incidence and prevalence of the condition, it is important to acknowledge that it may not necessarily align with the true incidence and prevalence due to various factors. While the data source itself may be reliable, it is worth noting that insurance claims for a specific diagnosis are often based on initial impressions and medical history. This can introduce potential inaccuracies, as these impressions may not always align with confirmed diagnoses by electrophysiologists or other specialists. Therefore, caution should be exercised when interpreting the data, as it may not fully capture the true prevalence and incidence of the condition.

## Conclusions

A correlation exists between the number of patients diagnosed with atrial fibrillation or flutter and average wind speed (negative) as well as average sunshine duration (positive) in the general population, with stronger associations observed in males aged over 35 and females aged over 55. The correlation with intraday temperature variation varies across different age and sex groups and is not consistent. Among females aged 20 to 24 years, there is a correlation with average temperature, precipitation, humidity, and atmospheric pressure. However, it is crucial to note that these correlations are specific to particular regions and should not be interpreted as indicating a causal relationship.

## Data Availability

Data is available upon request or can be accessed online through opendata.hira.or.kr, kma.go.kr, and kosis.kr.

kosis.kr

kma.go.kr

opendata.hira.or.kr

## Duplicate Publication

The abstract of this publication was presented at the Korean Heart Rhythm Society conference in Seoul on June 22, 2023, where it received a Young Investigator Award.

## Supplemental Materials

***(These original data are provided for review purposes only, but can be made available upon request by readers.)***

Supplemental 1: Total Population Statistics

Supplemental 2: Atrial Fibrillation and Flutter Statistics

Supplemental 3: Climate Statistics

Supplemental 4: Air Pollution Statistics

Supplemental 5-1: Pearson Correlation

Supplemental 5-2: Pearson Correlation (Colored)

## Clinical Trial Registration

*This is not a clinical trial*.

